# Maximum likelihood perimetric progression analysis: Using raw (trial-by-trial) response data to estimate progression more robustly

**DOI:** 10.1101/2021.02.05.21251210

**Authors:** Pete R. Jones

## Abstract

**Purpose:** To describe and demonstrate a more efficient (Maximum Likelihood) method for quantifying visual field progression.

**Design:** Monte Carlo simulation.

**Methods:** Trial-by-trial response data were simulated using a stochastic psychometric model (a “simulated observer”). Simulated Differential Light Sensitivity (DLS) decreased between tests to mimic long-term visual field progression. Progression slopes were fitted, either by fitting a regression slope to independent DLS estimates from each test (conventional method), or by fitting all the raw data combined in a single model (proposed maximum likelihood method).

**Results:** The proposed ML method seldom performed worse than a conventional, regression-based approach, and often performed better. For an idealized observer with a lapse (false negative) rate of 0 and a guess (false positive) rate of 0, both methods were equally precise. However, as lapse rate increased, the ML method exhibited less random measurement error. For small numbers of trials this increase in precision translated to a negative progression slope being detected with 95% confidence at least one year/assessment sooner. The only time the ML method was observed to perform worse was when very few trials (*N* = 4) were combined with very high lapse rates (λ = 0.3): an unlikely but not inconceivable scenario.

**Conclusions:** Combining raw, trial-by-trial response data in a single ML model can provide a more robust estimate of visual field progression than conventional methods (e.g., linear regression), at no additional cost to the patient or clinician (i.e., no additional trials).

---

The ability to accurately monitor perimetric rate of change (‘progression’) is key for the effective management of glaucoma. Typically, rate of progression (e.g., change in Differential Light Sensitivity, DLS, at a specific retinal location, or in some overall summary metric, such as Mean Deviation, MD) is estimated by fitting a curve through a series of point-estimates (scalar ‘best guess’ values), with each value representing a single assessment/hospital-visit. This approach is suboptimal, however, since there is additional information contained in the raw, trial-by-trial, test data; information that is effectively discarded when a point-estimate is generated at the end of each assessment.

In principle, this additional information could also be factored in when estimating rate of progression. Thus, rather than using the raw data from each test to compute independent point-estimates, ⍰DLS_0_, DLS_1_, …, DLS_n_⍰, and then fitting a line through this time-series (a ‘two step’ solution, illustrated in Fig 1A), all of the raw data from every test could instead be used together to fit a single, maximum likelihood (ML) model, consisting of a single psychometric function whose threshold is determined by two parameters: an initial value, DLS_0_, and a rate of progression value, DLS_Δ_ (a ‘one step’ solution, illustrated in Fig 1B). MATLAB code instantiating both of these methods is given as ***Supplemental Code***, and this same code was used to generate the simulations reported below.

**Figure 1.**
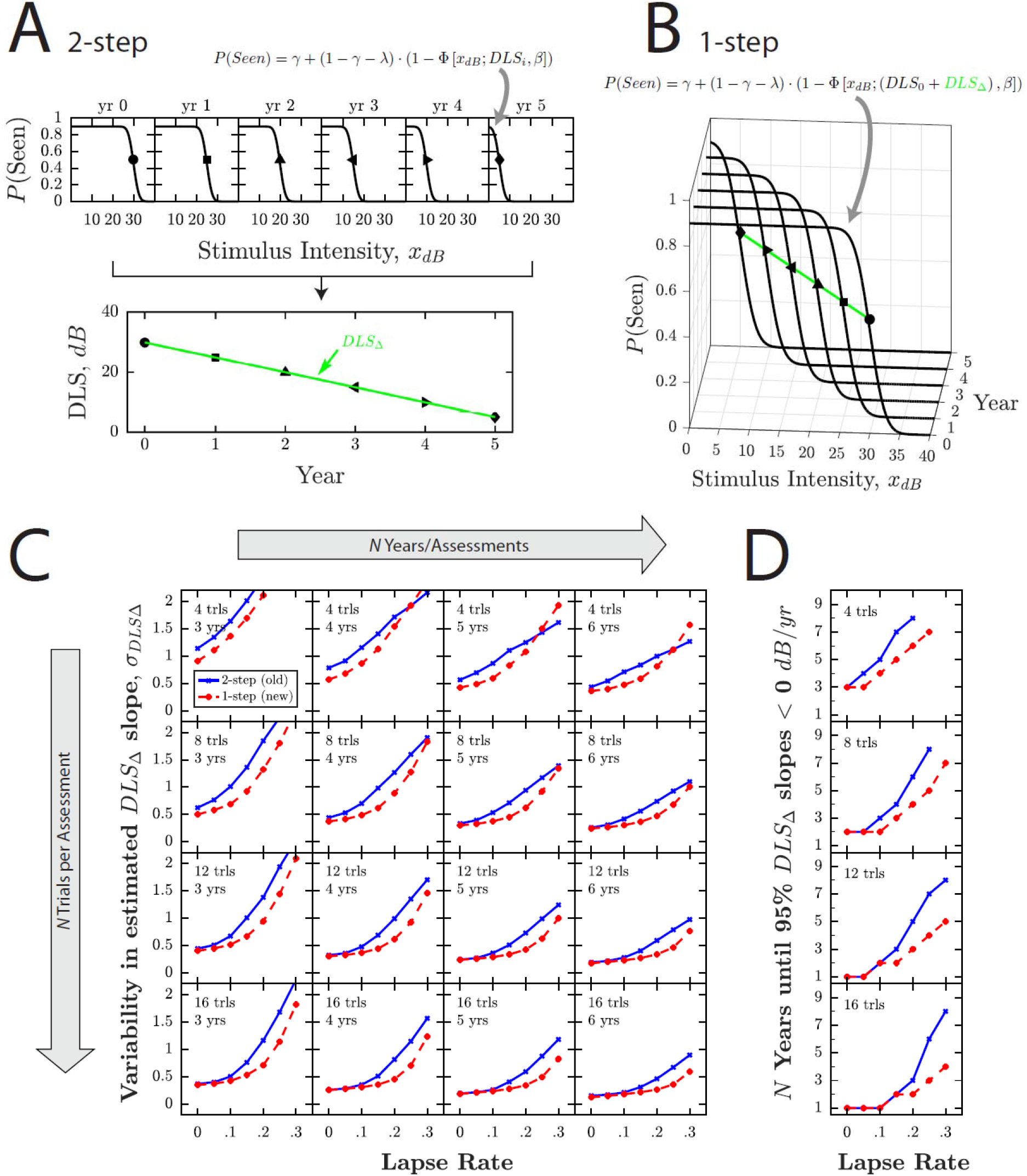
Methods and Results. ***(A)*** Schematic illustrations of conventional progression analysis: a two-step approach, in which DLS thresholds are first estimated independently for each assessment (e.g., using a maximum likelihood algorithm or adaptive staircase), and then a curve is fitted to this series of point-estimates (e.g., a least-squares linear regression slope). ***(B)*** Our proposed maximum likelihood method: a one-step approach in which the raw, trial-by-trial data from every test are used together to fit a single psychometric function, the threshold/location parameter of which, DLS_dB_, is determined by a starting value, DLS_S_, and rate of progression value, DLS_Δ_. ***(C)*** Simulation Results: variability in estimated rate of change, σ_DLSΔ,_ for the conventional method (blue crosses) and proposed ML method (red circles), as a function of lapse rate. Each datapoint represents 10,000 simulations (2.8M simulations in total), with a given *N Trials* per assessment, ⍰4, 8, 12, or 16 trials⍰, *N Assessments*, ⍰3, 4, …, or 12 years⍰, and *Lapse Rate*, ⍰0.05, 0.10, …, 0.30⍰. ***(D)*** Further analysis. Same underlying data as (C), but here showing the number of years until 95% of simulations (i.e., 9,500 simulations) produced a negative slope estimate: DLS_Δ_ < 0. This can be thought of as the number of years until the negative progression slope was detected with 95% confidence.

To assess whether this ‘one step’ (ML) solution has the potential to meaningfully reduce measurement error, we ran Monte Carlo simulations using a simulated observer, whose probability of responding correctly to a stimulus of magnitude dB was determined, in the typical fashion, by a psychometric function of the form:

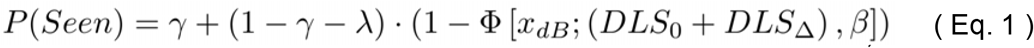

where Φ is a Gaussian cumulative density function (CDF); *DLS*_0_ is the initial threshold sensitivity at first assessment, fixed at 30 dB; DLS_Δ_ is the rate of progression, fixed at −1 dB/year (i.e., DLS = 25 dB after 5 years; Note that the sum of *DLS*_0_ and *DLS*_Δ_ together constituted the mean/location parameter of the CDF); γ is the guess rate (0 to 1), fixed at 0; *β* is the psychometric slope parameter (the standard deviation of the CDF), fixed at 2 dB (Note, not be confused with the rate of progression slope parameter: *DLS*_Δ_); and λ is the lapse rate (0 to 1), which was systematically manipulated across simulations as follows: ⍰0, 0.05, …, 0.3⍰.

Each simulated observer was used to generate ⍰3, 4, …, or 12⍰ ‘years’ of DLS estimates (1 assessment per year), with each assessment consisting of a single DLS estimate (a ‘1 point grid’) made using either ⍰8, 16, 32, or 64⍰ trials. This resulted in 280 unique conditions (7 *Lapse Rates X 10 N Years/Assessments X 4 N Trials Per Assessment*). Each of these 280 conditions was independently simulated 10,000 times, yielding 2.8M simulated ‘patients’ in total (each of whom performed 3—12 DLS assessments). Within each assessment, stimulus selection was determined by a conventional QUEST+ routine^1^ (in this instance equivalent to ZEST^2^), using a bimodal prior taken from Vingrys & Pianta (1999) ^3^. The final threshold estimate (i.e., *DLS*, in dB) was computed as the mean of the probability mass function. See ***Supplemental Code*** for full technical details.

For each simulated patient, rate of progression was then computed, post hoc, in two ways: First, using a conventional two-step linear regression procedure (***Fig 1A***). Second, by refitting all of the raw, trial-by-trial data from every assessment using the proposed ML procedure (***Fig 1B***). This ML procedure differed from the conventional QUEST+/ZEST procedure used within each individual assessment only inasmuch as the stimulus domain had two dimensions (*Stimulus Intensity* in dB, and *N Years*) rather than one (*Stimulus Intensity*) and the model free-parameters domain had two dimensions (*DLS*_0_ and *DLS*_Δ_) rather than one (*DLS*).

For each set of 10,000 rate of progression estimates, the standard deviation, σ_DLSΔ_, was computed as an index of random measurement error. Ideally, σ_DLSΔ_ would be 0 (same estimated rate of progression on every simulation), with higher values indicating lower precision.

The results are shown in **Figure 1C,D**. In brief, the proposed ML method seldom performed worse than a conventional, regression-based approach, and often performed better. For an idealized observer with a lapse (false negative) rate of 0 and a guess (false positive) rate of 0, both methods were equally precise (**Fig 1C**, *leftmost data point in each panel*). However, as lapse rate increased, the ML method exhibited less random measurement error. For small numbers of trials this increase in precision translated to a negative progression slope being detected with 95% confidence at least one year/assessment sooner (**Fig 1D**). Unsurprisingly, this difference between the two methods was attenuated when large, clinically unrealistic, numbers of trials were used to estimate thresholds, since in such circumstances measurement error for both methods converged towards zero (**Fig 1C,D**, *bottommost panels*). Though some advantage for the ML method was still observed for very high lapse rates. The only time the ML method was observed to perform more poorly than the conventional method was when very few trials (N = 4) were combined with very high lapse rates (λ = 0.3): an unlikely but not inconceivable scenario.

Other than the data shown in **Figure 1**, no other conditions (i.e., observer parameters or psychometric algorithm settings) were simulated: The goal was not to systematically assess the exact level of benefit under all possible scenarios, but to assess whether any meaningful benefit is possible under a single, broadly realistic scenario. Interested readers can, however, modify the ***Supplemental Code*** to simulate other scenarios.

These simulations, though not comprehensive, illustrate that combining raw, trial-by-trial response data in a single ML model can provide a more robust estimate of visual field progression than a conventional ‘two-step’ (e.g., linear regression) approach, at no additional cost to the patient or clinician (i.e., without requiring any additional stimulus presentations, or varying current test durations or methods in any way).

## Supporting information

Supplemental MATLAB Code

## Data Availability

Source code provided as Supplemental Material

## Notes

**<u>FINANCIAL SUPPORT:</u>** ⟨ none ⟩

**<u>CONFLICTS OF INTEREST:</u>** No conflicting relationship exists for any author.

### Competing Interest Statement

The authors have declared no competing interest.

### Funding Statement

No funding to declare

## REFERENCES

1. Watson, A. B. QUEST+: A general multidimensional Bayesian adaptive psychometric methodWatson. J. Vis. 2017;17:10

2. Turpin, A., McKendrick, A. M., Johnson, C. A. & Vingrys, A. J. Properties of perimetric threshold estimates from full threshold, ZEST, and SITA-like strategies, as determined by computer simulation. Invest. Ophthalmol. Vis. Sci. 2003;44:4787–4795

3. Vingrys, A. J. & Pianta, M. J. A new look at threshold estimation algorithms for automated static perimetry. Optom. Vis. Sci. 1999;76:588–595

