## Supplemental MATLAB Code for "Maximum likelihood perimetric progression analysis: Using raw (trial-by-trial) response data to estimate progression more robustly": MLprogressionAnalysis_SuplCode.m

```
% Script for applying different forms of post-hoc progression analysis to
% stochastic data from a simulated observer. Tested in MATLAB 2016b only. 
%
% To reference, and for further info:
%	<a href="xxxx">Jones, P.R. (in submission). Maximum-likelihood perimetric progression analysis: Using raw, trial-by-trial response-data to estimate progression more robustly.</a>
%
% Dependencies:
%   QuestPlus.m  => download from https://github.com/petejonze/QuestPlus
%   fig-matlab toolbox (if doPlot==true) => download from https://github.com/petejonze/fig-matlab
%   MATLAB Statistics Toolbox (though could remove this dependency by replacing "regstats()")
%
% Author:
%   Pete R Jones <>
%
% Verinfo:
%   1.0     PJ  12/2020 : first_build\n
%
% This code is licenced under CC BY-SA 3.0. This permits the code to be
% used for free, even in commercial applications, as long as: (1) the
% present work is cited, and (2) the derivative work is licensed under the
% same terms.
%
% Copyright 2020 : P R Jones
% *********************************************************************
% 

% init
clc
close all

% User parameters
verbosity       = 0;
TRUE_LAPSE_RATE = 0:.05:.3;
TRUE_GUESS_RATE = 0.00 * ones(size(TRUE_LAPSE_RATE));
nTRIALS         = [4 8 12 16];
nYEARS          = 3:10;
N_SIMULATIONS   = 10000; % 10000; % 200
doPlot          = true;
saveFigs      	= true;


% *************************************************************************
% -------------------- NO CHANGES BEYOUND THIS POINT -------------------- %
% *************************************************************************


%%%%%%%
%% 1 %%
%%%%%%%%%%%%%%%%%%%%%%%%%%%%%%%%%%%%%%%%%%%%%%%%%%%%%%%%%%%%%%%%%%%%%%%%%%%
%%% Initialise                                                          %%%
%%%%%%%%%%%%%%%%%%%%%%%%%%%%%%%%%%%%%%%%%%%%%%%%%%%%%%%%%%%%%%%%%%%%%%%%%%%

% initialise figure plotting
[EXPORT_DIR, EXPORT_FORMAT] = fig_init(0.3, 'pdf', []); % though any heatmaps will be PNG
if ~saveFigs
    EXPORT_DIR = NaN; % uncomment to suppress figure saving
end

% compute further variables
N_LAPSE_GUESS_RATE_COMBINATIONS = length(TRUE_LAPSE_RATE);
N_nTRIALS = length(nTRIALS);
N_nYEARS = length(nYEARS);

% init priors: priors taken from population dat (Vingrys & Pianta, Optom &
% Vis Sci, 1999: "A new look at...")
normativePriors = [ 
    0    0.0285
    1.0000    0.0238
    2.0000    0.0194
    3.0000    0.0155
    4.0000    0.0124
    5.0000    0.0102
    6.0000    0.0082
    7.0000    0.0062
    8.0000    0.0053
    9.0000    0.0045
    10.0000    0.0038
    11.0000    0.0035
    12.0000    0.0035
    13.0000    0.0038
    14.0000    0.0045
    15.0000    0.0053
    16.0000    0.0064
    17.0000    0.0074
    18.0000    0.0090
    19.0000    0.0105
    20.0000    0.0127
    21.0000    0.0152
    22.0000    0.0193
    23.0000    0.0243
    24.0000    0.0304
    25.0000    0.0387
    26.0000    0.0507
    27.0000    0.0654
    28.0000    0.0819
    29.0000    0.0948
    30.0000    0.0957
    31.0000    0.0849
    32.0000    0.0642
    33.0000    0.0451
    34.0000    0.0309
    35.0000    0.0206
    36.0000    0.0130
    37.0000    0.0087
    38.0000    0.0057
    39.0000    0.0037
    40.0000    0.0025];


%%%%%%%
%% 2 %%
%%%%%%%%%%%%%%%%%%%%%%%%%%%%%%%%%%%%%%%%%%%%%%%%%%%%%%%%%%%%%%%%%%%%%%%%%%%
%%% Run                                                                 %%%
%%%%%%%%%%%%%%%%%%%%%%%%%%%%%%%%%%%%%%%%%%%%%%%%%%%%%%%%%%%%%%%%%%%%%%%%%%%

for USE_PRIORS = [0 2]
    
    % init output variables
    slopeEst_QP = nan(N_SIMULATIONS, N_nTRIALS, N_nYEARS, N_LAPSE_GUESS_RATE_COMBINATIONS);
    slopeEst_Reg = nan(N_SIMULATIONS, N_nTRIALS, N_nYEARS, N_LAPSE_GUESS_RATE_COMBINATIONS);
    
    % prepare to run
    fprintf('Running simulations\n');
    tic();
    if verbosity == 0
        warning off;
    end
    
    for i0 = 1:N_nTRIALS
        fprintf(' .');
        for i1 = 1:N_nYEARS
            for i2 = 1:N_LAPSE_GUESS_RATE_COMBINATIONS
                for k = 1:N_SIMULATIONS
                    
                    years = 0:nYEARS(i1);
                    all_history_stim = [];
                    all_history_resp = [];
                    all_history_year = [];
                    mu_est_each_year = nan(size(years));
                    
                    for i = 1:length(years)
                        year = years(i);
                        
                        if verbosity>0
                            fprintf('\n\n**************************************** Year %i ****************************************\n', year);
                        end
                        
                        % define model: Probability of responding correctly to a given stimulus
                        % magnitude (magnitude in conventional SAP dB scale, so bigger values are
                        % HARDER to see)
                        F = @(stimLvl_dB,mu,sigma,gamma,lambda)(gamma+(1-gamma-lambda).*(1-normcdf(stimLvl_dB,mu,sigma)));
                        
                        % set true params of our simulated observer [for simulation purposes only]
                        pf_mu_baseline	= 30; % 20;                   % psychometric function: mean
                        pf_mu_delta    	= -1;                   % progression: rate at which mu changes per year
                        pf_beta         = 2;                    % psychometric function: slope
                        pf_gamma        = TRUE_GUESS_RATE(i2);	% psychometric function: guess rate
                        pf_lambda       = TRUE_LAPSE_RATE(i2);  % psychometric function: lapse rate
                        
                        % define stimulus domain
                        stimLvl_dB = linspace(0,40,41);
                        stimDomain = {stimLvl_dB};
                        
                        % define parameter domain
                        mu_dB    	= linspace(0,40,41);
                        beta_dB    	= 2;
                        gamma_0to1	= 0.1;
                        lambda_0to1	= 0.1;
                        paramDomain = {mu_dB, beta_dB, gamma_0to1, lambda_0to1};
                        
                        % define response domain
                        respDomain = [0 1];
                        
                        % define priors
                        if USE_PRIORS>0
                            priors = {
                                normativePriors(:,2)./sum(normativePriors(:,2))
                                1
                                1
                                1
                                };
                        else
                            priors = []; % will assume uniform for all
                        end
                        
                        % define QUEST+ stop rule parameters (blank for default)
                        stopRule        = [];
                        stopCriterion   = [];
                        minNTrials      = nTRIALS(i0);
                        maxNTrials      = nTRIALS(i0);
                        
                        % create QUEST+ object
                        QP = QuestPlus(F, stimDomain, paramDomain, respDomain, stopRule, stopCriterion, minNTrials, maxNTrials);
                        
                        % initialise priors/likelihoods
                        fn = 'likelihoods_singleSession.mat';
                        if exist(fn, 'file')
                            QP.initialise(priors, fn, verbosity);
                        else
                            QP.initialise(priors);
                            QP.saveLikelihoods(fn);
                        end
                        
                        % get starting parameter estimates
                        startGuess_mean = QP.getParamEsts('mean');
                        
                        % display starting estimates
                        if verbosity>0
                            fprintf('========================================\nSTARTING ESTIMATES\n====================\n');
                            %QP.disp();
                            fprintf('mu_dB estimate:        %7.2f	[true: %1.2f]\n', startGuess_mean(1), pf_mu_baseline + pf_mu_delta*year);
                            fprintf('beta_dB estimate:      %7.2f	[true: %1.2f]\n', startGuess_mean(2), pf_beta);
                            fprintf('gamma_0to1 estimate:   %7.2f	[true: %1.2f]\n', startGuess_mean(3), pf_gamma);
                            fprintf('lambda_0to1 estimate:	%7.2f	[true: %1.2f]\n', startGuess_mean(4), pf_lambda);
                        end
                        
                        % run -------------------------------------------------
                        while ~QP.isFinished()
                            stimLvl_dB = QP.getTargetStim();
                            pC = F(stimLvl_dB, pf_mu_baseline + pf_mu_delta*year, pf_beta, pf_gamma, pf_lambda);
                            anscorrect = rand() < pC;
                            QP.update(stimLvl_dB, anscorrect);
                        end
                        
                        % get final parameter estimates
                        endGuess_mean = QP.getParamEsts('mean');
                        
                        % display final estimates
                        if verbosity>0
                            fprintf('========================================\nFINAL ESTIMATES\n====================\n');
                            %QP.disp();
                            fprintf('mu_dB estimate:        %7.2f	[true: %1.2f]\n', endGuess_mean(1), pf_mu_baseline + pf_mu_delta*year);
                            fprintf('beta_dB estimate:      %7.2f	[true: %1.2f]\n', endGuess_mean(2), pf_beta);
                            fprintf('gamma_0to1 estimate:   %7.2f	[true: %1.2f]\n', endGuess_mean(3), pf_gamma);
                            fprintf('lambda_0to1 estimate:	%7.2f	[true: %1.2f]\n', endGuess_mean(4), pf_lambda);
                        end
                        
                        % store
                        all_history_stim    = [all_history_stim, QP.history_stim]; %#ok<AGROW>
                        all_history_resp    = [all_history_resp, QP.history_resp]; %#ok<AGROW>
                        all_history_year    = [all_history_year, year*ones(1,length(QP.history_resp))]; %#ok<AGROW>
                        mu_est_each_year(i) = endGuess_mean(1);
                    end
                    
                    
                    % ================================================
                    if verbosity>0
                        fprintf('\n\n**************************************** META ****************************************\n');
                    end
                    
                    % define model
                    F_meta = @(stimLvl_dB,nYearsSince1stTest, mu,mu_delta,sigma,gamma,lambda)(gamma+(1 - gamma - lambda).*(1-normcdf(stimLvl_dB,mu + mu_delta*nYearsSince1stTest,sigma)));
                    
                    % define stimulus domain
                    stimLvl_dB = linspace(0,40,41);
                    year       = 0:max(nYEARS);
                    stimDomain = {stimLvl_dB, year};
                    
                    % define parameter domain
                    mu_baseline_dB      = linspace(0,40,41);
                    mu_delta_dBPerYr	= linspace(-10,10,41); % linspace(-10,0,21);
                    beta_dB             = 2;
                    gamma_0to1          = .1; % [0 0.05 .1 .2];
                    lambda_0to1         = .1; % [0 0.05 .1 .2];
                    paramDomain = {mu_baseline_dB, mu_delta_dBPerYr, beta_dB, gamma_0to1, lambda_0to1};
                    
                    % define response domain
                    respDomain = [0 1];
                    
                    % define priors
                    if USE_PRIORS>1
                        priors = {
                            (normativePriors(:,2)./sum(normativePriors(:,2)))'
                            ones(1,41)./41
                            1
                            1
                            1
                            };
                    else
                        priors = []; % will assume uniform for all
                    end
                    
                    % create QUEST+ object
                    QP = QuestPlus(F_meta, stimDomain, paramDomain, respDomain);
                    
                    % initialise priors/likelihoods
                    fn = 'likelihoods_meta.mat';
                    if exist(fn, 'file')
                        QP.initialise(priors, fn, verbosity);
                    else
                        QP.initialise(priors);
                        QP.saveLikelihoods(fn);
                    end
                    
                    % get starting parameter estimates
                    startGuess_mean = QP.getParamEsts('mean');
                    
                    % % display starting estimates
                    if verbosity>0
                        fprintf('========================================\nSTARTING ESTIMATES\n====================\n');
                        %QP.disp();
                        fprintf('mu_baseline_dB estimate:   %7.2f	[true: %1.2f]\n', startGuess_mean(1), pf_mu_baseline);
                        fprintf('mu_delta_dBPerYr estimate: %7.2f 	[true: %1.2f]\n', startGuess_mean(2), pf_mu_delta);
                        fprintf('beta_dB estimate:          %7.2f	[true: %1.2f]\n', startGuess_mean(3), pf_beta);
                        fprintf('gamma_0to1 estimate:       %7.2f	[true: %1.2f]\n', startGuess_mean(4), pf_gamma);
                        fprintf('lambda_0to1 estimate:      %7.2f	[true: %1.2f]\n', startGuess_mean(5), pf_lambda);
                    end
                    
                    % run -------------------------------------------------
                    for i = 1:length(all_history_stim)
                        stimLvl_dB  = all_history_stim(i);
                        year        = all_history_year(i);
                        anscorrect  = all_history_resp(i);
                        QP.update([stimLvl_dB; year], anscorrect);
                    end
                    
                    % get final parameter estimates
                    endGuess_mean = QP.getParamEsts('mean');
                    
                    % display final estimates
                    if verbosity>0
                        fprintf('========================================\nFINAL ESTIMATES\n====================\n');
                        %QP.disp();
                        fprintf('mu_baseline_dB estimate:   %7.2f	[true: %1.2f]\n', endGuess_mean(1), pf_mu_baseline);
                        fprintf('mu_delta_dBPerYr estimate: %7.2f 	[true: %1.2f]\n', endGuess_mean(2), pf_mu_delta);
                        fprintf('beta_dB estimate:          %7.2f	[true: %1.2f]\n', endGuess_mean(3), pf_beta);
                        fprintf('gamma_0to1 estimate:       %7.2f	[true: %1.2f]\n', endGuess_mean(4), pf_gamma);
                        fprintf('lambda_0to1 estimate:      %7.2f	[true: %1.2f]\n', endGuess_mean(5), pf_lambda);
                    end
                    
                    % crude regression model
                    stats = regstats(mu_est_each_year, years);
                    
                    % STORE
                    slopeEst_Reg(k, i0, i1, i2) = stats.beta(2);
                    slopeEst_QP(k, i0, i1, i2) = endGuess_mean(2);
                end
            end
        end
    end
    
    if verbosity == 0
        warning on;
    end
    
    fprintf('\nFinished simulations\n');
    toc()
    fprintf('\n\n');
    
    %% save data
    save(sprintf('data_nSims=%i_UsePriors=%i_%s.mat',N_SIMULATIONS,USE_PRIORS,datestr(now(),30)));
    
    %% compute N Years until change detected
    prcnt = 95;
    nYrs_Reg = nan(N_nTRIALS,length(TRUE_LAPSE_RATE));
    nYrs_QP = nan(N_nTRIALS,length(TRUE_LAPSE_RATE));
    
    for i = 1:N_nTRIALS
        
        for cid = 1:length(TRUE_LAPSE_RATE)
            idx_cond = TRUE_LAPSE_RATE==TRUE_LAPSE_RATE(cid) & TRUE_GUESS_RATE==TRUE_GUESS_RATE(cid);
            idx_nTrials = nTRIALS==nTRIALS(i);
            
            % 2 step
            tmp = find(squeeze(prctile(slopeEst_Reg(:,idx_nTrials,:,idx_cond),prcnt))<0,1,'first'); %todo: use bootstrapping to derive CI
            if ~isempty(tmp)
                nYrs_Reg(i,cid) = tmp;
            end
            
            % 1 step
            tmp = find(squeeze(prctile(slopeEst_QP(:,idx_nTrials,:,idx_cond),prcnt))<0,1,'first');
            if ~isempty(tmp)
                nYrs_QP(i,cid) = tmp;
            end
        end
    end
    
    %% Lattice: plot SD as a function of lapse rate, for different nTrials (rows) and different nYears (columns)
    if doPlot
        nYEARS1 = [3 4 5 6]; % [3 5 8 12];
        N_nYEARS1 = length(nYEARS1);
        
        % open a new figure window
        figDim      = [20 20];
        nPlots      = [N_nTRIALS N_nYEARS1+1];
        isTight     = true;
        isSquare    = true;
        styleFlag   = [];
        axesLims    = [];
        hFig        = fig_make(figDim, nPlots, isTight, isSquare, styleFlag, axesLims);
        
        for i = 1:N_nTRIALS
            fig_subplot(i,N_nYEARS1+1);
            fig_nudge(gca,.08,0);
            hDat = plot(TRUE_LAPSE_RATE,nYrs_Reg(i,:),'bx-', TRUE_LAPSE_RATE,nYrs_QP(i,:),'r*--');
            set(hDat,'linewidth',1.5, 'markersize',4);
            set(gca,'ytick',[1 2 3],'yticklabel',[1 2 3]);
            
            % for debugging
            txt = sprintf('%i trls',nTRIALS(i));
            textLoc(txt, 'NorthWest');
        end
        
        % plot left lattice (variability)
        for i = 1:N_nTRIALS
            for j = 1:N_nYEARS1
                
                fig_subplot(i,j);
                hold on
                idx_nTrials = nTRIALS==nTRIALS(i);
                idx_nYears = nYEARS==nYEARS1(j);
                
                hDat = plot(TRUE_LAPSE_RATE,squeeze(std(slopeEst_Reg(:, idx_nTrials, idx_nYears, :))),'bx-', TRUE_LAPSE_RATE,squeeze(std(slopeEst_QP(:, idx_nTrials, idx_nYears, :))),'r*--');
                set(hDat,'linewidth',1.5, 'markersize',4);
                
                % for debugging
                %txt = sprintf('%i trials, %i yrs',nTRIALS(i),nYEARS1(j));
                txt = {sprintf('%i trls',nTRIALS(i)), sprintf('%i yrs',nYEARS1(j))};
                textLoc(txt, 'NorthWest');
                
                % format (all) the axes
                xTick       = 0:.1:.3;
                xTickLabels = {'0','.1','.2','.3'};
                yTick       = 0:.5:2;
                yTickLabels = [];
                xTitle      = [];
                yTitle      = [];
                xlims       = [-.05 0.35];
                ylims       = [-.2 2.2];
                fontSize    = [16 11];
                fig_axesFormat(gca, xTick,xTickLabels, yTick,yTickLabels, xTitle,yTitle, xlims,ylims, fontSize);
            end
        end
        
        % plot right lattice (N years until 95%)
        for i = 1:N_nTRIALS
            fig_subplot(i,N_nYEARS1+1);
            
            yTick       = 1:2:9;
            yTickLabels = [];
            xTitle      = [];
            yTitle      = [];
            xlims       = [-.05 0.35];
            ylims       = [0.5 10];
            fontSize    = [16 11];
            formatData	= [];
            xMinorTick  = [];
            yMinorTick  = 2:2:8;
            mrTickLgth  = .66;
            fig_axesFormat(gca, xTick,xTickLabels, yTick,yTickLabels, xTitle,yTitle, xlims,ylims, fontSize, formatData, xMinorTick,yMinorTick,mrTickLgth);
        end
        
        % format the figure
        xTitle = '\textbf{Lapse Rate}';
        yTitle = '\textbf{Variability in estimated slope}, $\sigma_{\Delta}$'; % '\textbf{ECL $P$(Hit)}';
        fontSize = 16;
        [hXTitle,hYTitle] = fig_figFormat(hFig, xTitle,yTitle, [], fontSize);
        fig_nudge(hXTitle,-2,0);
        
        % add axis labels to right plot too
        hXTitle2 = copyobj(hXTitle,hXTitle.Parent);
        fig_nudge(hXTitle2,9,0);
        hYTitle2 = copyobj(hYTitle,hYTitle.Parent);
        hYTitle2.String = '\textbf{$N$ Years until 95\% slopes $<$ 0 $dB/yr$}';
        fig_nudge(hYTitle2,14.6,0);
        
        % add legend
        fig_subplot(1)
        hLeg = legend('Old','New','Location','SouthEast');
        set(hLeg,'interpreter','latex');
        hLeg.ItemTokenSize(1) = 18;
        
        % save
        fn = sprintf('results_v1_nSims=%i_UsePriors=%i_%s',N_SIMULATIONS,USE_PRIORS,datestr(now(),30));
        fig_save(hFig, fn, EXPORT_DIR, EXPORT_FORMAT);
    end
end
```
